# Altered Resting State Brain Networks and Cognition in Familial Adenomatous Polyposis

**DOI:** 10.1101/2020.11.02.20224477

**Authors:** Ryan J. Cali, Benjamin C. Nephew, Constance M. Moore, Serhiy Chumachenko, Ana Cecilia Sala, Beatriz Cintron, Carlos Luciano, Jean A. King, Stephen R. Hooper, Francis M. Giardiello, Marcia Cruz-Correa

## Abstract

Familial Adenomatous Polyposis (FAP) is an autosomal dominant disorder caused by mutation of the APC gene presenting with numerous colorectal adenomatous polyps and a near 100% risk of colon cancer. Preliminary research findings from our group indicate that FAP patients experience significant deficits across many cognitive domains. In the current study, fMRI brain metrics in a FAP population and matched controls were used to further the mechanistic understanding of reported cognitive deficits. This research identified and characterized any possible differences in resting brain networks and associations between neural network changes and cognition from 34 participants (18 FAP patients, 16 healthy controls). Functional connectivity analysis was performed using FSL with independent component analysis (ICA) to identify functional networks. Significant differences between cases and controls were observed in 8 well-established resting state networks. With the addition of an aggregate cognitive measure as a covariate, these differences were virtually non-existent, indicating a strong correlation between cognition and brain activity at the network level. The data indicate robust and pervasive effects on functional neural network activity among FAP patients and these effects are likely involved in cognitive deficits associated with this disease.

## Introduction

Familial Adenomatous Polyposis (FAP) is an autosomal dominant disorder caused by germline mutations in the APC (adenomatous polyposis coli) gene. This disease presents at young age with multiple adenomatous polyps in the colorectum and leads to early-onset cancer. If left untreated, these patient develop colorectal on average by the fourth decade of life (Fearnhead, Britton, & Bodmer, 2001). The frequency of the disease is estimated to be 1 in 8,000 persons (Bisgaard, Fenger, Bulow, Niebuhr, & Mohr, 1994).

Based on anecdotal clinical observations of behavioral and cognitive difficulties, researchers have recently become interested in examining cognitive function in FAP patients. A 2010 study by researchers at the Cleveland Clinic reported that though FAP patients have IQs parallel with the general population, hearing/language-dependent verbal scores were significantly lower than those without the mutation (O’Malley et al., 2010). Preliminary data in a small sample, sibling-paired pilot study suggested that siblings who are FAP positive are more likely to suffer from behavioral and emotional problems than their healthy siblings (Azofra et al., 2016), and a prior study reported a high incidence of formal psychiatric diagnoses (especially anxiety diagnoses) in FAP adolescents (Gjone, Diseth, Fausa, Nøvik, & Heiberg, 2011). The most recent investigation of neurocognitive function of FAP patients identified and characterized robust deficits in several cognitive measures (particularly long term retrieval and cognitive fluency), suggesting that APC protein plays a critical role in cognition (M. R. Cruz-Correa et al., 2020).

The pilot study from 2016 identified no significant differences in brain morphology comparing healthy siblings to FAP sibling counterparts, and this was the first time an FAP population had been studied for neuroanatomical differences using magnetic resonance imaging (MRI) (Azofra et al., 2016). Given lack of understanding of the neurological and neurocognitive effects of this disease, the objective of the current study was to develop a more robust understanding of how FAP affects brain function. The present study employed resting state functional MRI (fMRI) to investigate potential neural substrates for cognitive deficits in a homogenous Hispanic population. To our knowledge, this is the first study of FAP patients using resting-state fMRI, which is ideally suited to the study of neurocognitive differences and related mechanisms (Hawes, Sokolowski, Ononye, & Ansari, 2019; LaClair et al., 2019; Li & King, 2019; Picó-Pérez et al., 2019; Sripada et al., 2019).

## Methods

### MRI data acquisition

Magnetic resonance images were acquired using a GE Discovery MR750 3T Scanner (General Electric Healthcare, Chicago, Illinois). An axial resting-state functional magnetic resonance imaging scan was performed on 34 individuals, which included 18 with FAP and 16 healthy controls (HC). The group consisted of 16 males and 18 females, (mean age of controls: 30.75; cases: 31.44 years). Axial fMRI scan time was approximately 6:15 (mm/ss). The scanner was equipped with a standard 8-channel head coil. Scan parameters: Frequency FOV (22.4), Slice thickness (3.5), Flip angle (90), Pixel size (3.5×3.5), Pulse (Gradient Echo EPI), Sequential Slice Order, Acquisition TR (2500msec).

### Participants

Inclusion criteria for cases included genetically confirmed FAP (based on a mutation on the APC gene performed by commercial laboratory testing), age⍰≥⍰10⍰.years, able to assent (children) and consent (adults) to participate in this study, and able to complete all neurocognitive testing. Inclusion criteria for controls included no known family history of FAP, negative clinical diagnosis of FAP based on colonoscopy (for adults), and willingness to undergo neurocognitive testing. Participants were all Spanish-speaking individuals from Puerto Rico. As education and age are correlated with IQ, cases were matched to non-FAP control individuals with regards to age (± three years), gender, and education (less than high school, at least high school, bachelors-degree or post-graduate education). Exclusion criteria for both cases and controls included previous diagnosis of any major psychiatric condition given the potential impact of these conditions on neurocognitive functioning, inability to sign/assent/consent study participation, or complete the neurocognitive tests.

Participants signed informed consent (or assent for children) prior to participation in the study and were evaluated at the Puerto Rico Clinical and Translational Research Consortium at the University of Puerto Rico (UPR) Medical Sciences Campus. The research protocol was approved by the Institutional Review Board of the UPR Medical Sciences Campus.

### Independent Component Analysis (ICA)

Independent component analysis is a tool often used to separate a specific signal or “blind signal separation” from a host of signals (McKeown et al, 2010). In this study, ICA was employed to parse out the specific signals generated by fMRI and identify potential noise artifacts. Based on established practices in previous studies using FSL and ICA, a total number of 30 ICs were extracted to avoid overfitting and underfitting of the dataset (Woolrich et al, 2009). Group-level ICA was performed using well-defined resting state networks (RSNs)(Smith et al, 2009).

### Data Preprocessing

Raw fMRI images were processed using FSL (figure 2). Slice timing correction was implemented in a bottom/up order. Within FSL, Independent Component Analysis (ICA) was used to visualize significant components. Motion was corrected for by omitting any components that displayed rapid changes in both time series and power spectra after an initial pass using FSL’s MELODIC ICA tool with a subsequent second correction being done manually, removing any components that showed noise based on the recommendations of Griffanti et al, 2016. Images were then concatenated and dual regressed. FSL’s “randomize” function was implemented with a threshold of Z>2.3 or P<0.05 and 10,000 permutations were run to ensure significance held at the .05 level (n= 10,000, 0.0500 ± 0.0044). A matrix was designed within the tool to display any differences in rsFC when comparing control patients to cases (Control>Case, Case<Control). The networks presented in this paper were chosen based on commonly seen RSNs in previous studies that detailed network differences in populations with cognitive impairments.

**Table 1.** Demographic characterization and cognitive data of study population.

|  | **FAP (n = 18)** | **Controls (n=16)** |
| --- | --- | --- |
| **Male** | 8 | 8 |
| **Female** | 10 | 8 |
| **Age (years)** | 31.44 (SD 14.6 ± 3.44) | 30.75 (SD 15.22 ± 3.80) |
| **Cognitive Score (Batería III Woodcock Múñez)** | 78.50 (SD 12.04 ± 2.83) | 96 (SD 8.58 ± 2.14) |

**Figure 1:**
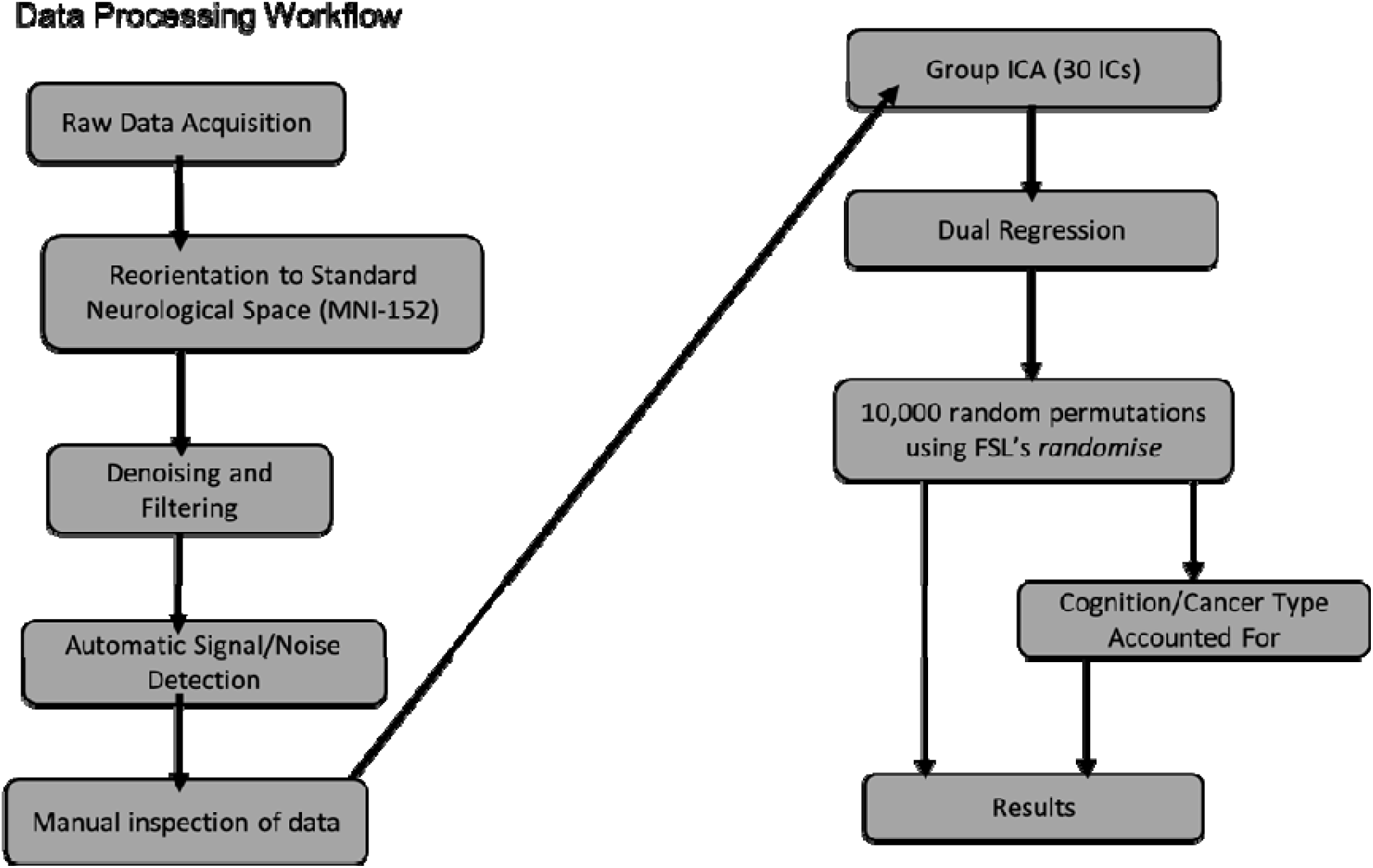
fMRI analysis workflow using Independent Component Analysis.

**Figure 2:**
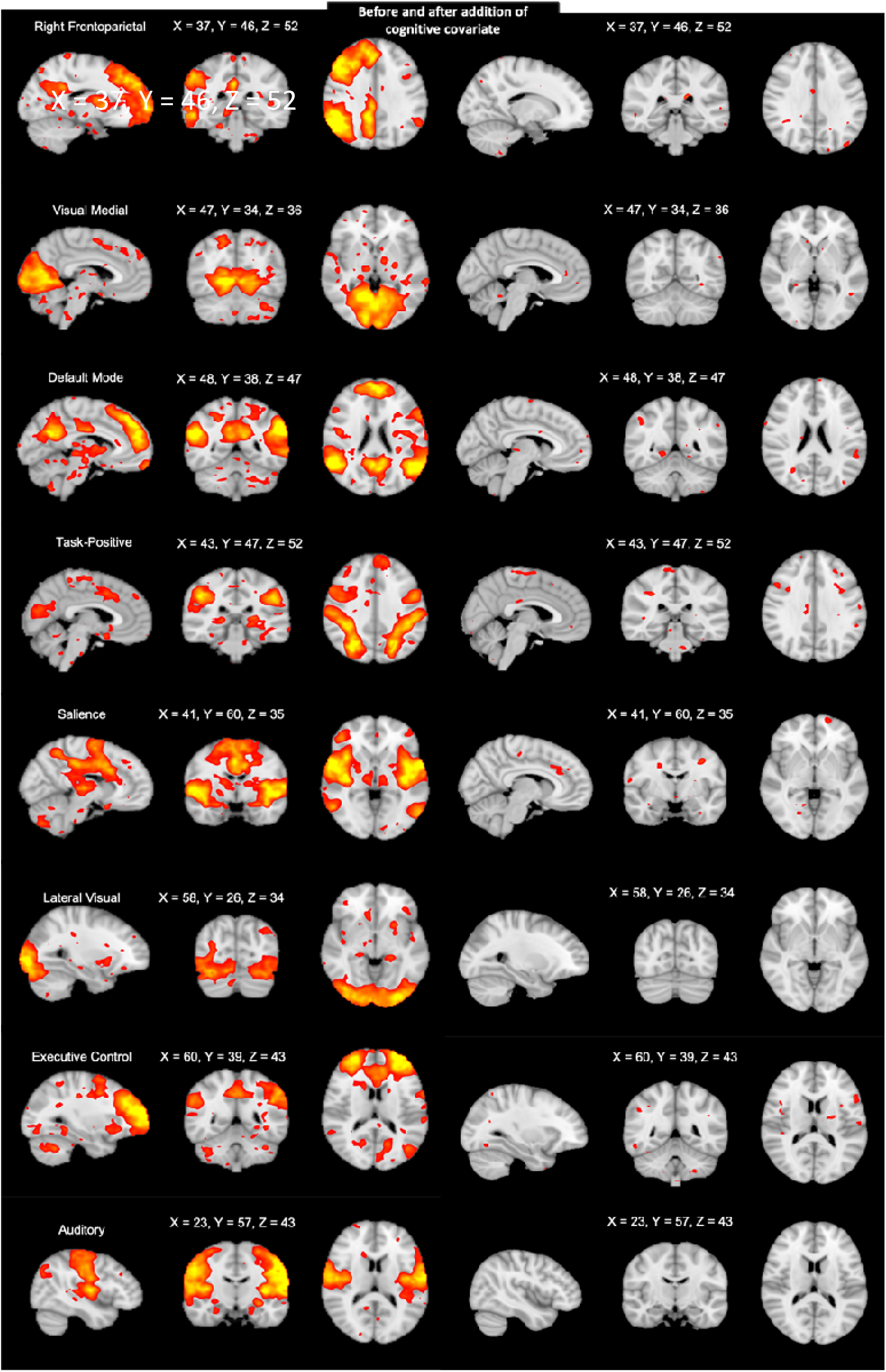
Activation maps of significant group differences projected onto MNI-152 standard space displaying clusters resembling 8 well-established brain networks (right, Smith et al, 2019). Upon introduction of the omnibus cognitive covariate into the GLM, cluster activation was no longer present (left).

### Cognitive Measures

Based on the cognitive assessments employed in the proof-of-concept paper (Cruz-Correa et al, 2020), an omnibus cognitive measure was created by averaging the nine cognitive domains from the Batería III Woodcock Munez. These measures consisted of: Cognitive Efficiency, Working Memory, Long-Term Retrieval, and Executive Function, Verbal Ability, Processing Speed, Phonemic Awareness, Cognitive Fluency, and Thinking Ability. This omnibus score was then averaged between groups and introduced as a covariate into the GLM matrix of the fMRI data. The FAP group had an average cognitive score of 78.50 (SD 12.04, N=18) and the mean for the healthy controls was 96 (SD 8.58, N= 16).

## Results

### Resting State

Differences in resting state data were seen in approximately 8 well-established networks identified in previous works (Smith et al., 2009). The Left Frontoparietal Network (LFPN), Visual Medial Network, Default Mode Network (DMN), Task-Positive Network (TPN), Salience Network, Lateral Visual Network, Executive Control Network (ECN), and Auditory Network were all noted as the most identifiable networks (fig. 1). rsfMRI data was coregistered to the standard MNI152 template for visualization (Montreal Neurological Institute) (Fonov et al., 2011). Additionally, a secondary analysis of the FAP group revealed no significant differences when accounting for genotype-phenotype severity using “attenuated” and “classic” FAP as covariate groupings (M. Cruz-Correa & Giardiello, 2003; M. R. Cruz-Correa et al., 2020).

### Cognition as a Covariate

Group differences between all nine of the cognitive variables were statistically significant both independently and when combined to form the aggregate score used in the rsfMRI analysis (p<0.05). When adjusting for cognition as a covariate to the rsfMRI data, significant differences were virtually eliminated, with only indications of minor differences throughout the components. This suggests a high correlation between cognitive functioning and rsfMRI findings.

## Discussion

The present study represents the first report of significant differences in resting state functional connectivity in FAP patients, with robust alterations in 8 brain networks which mediate cognition and sensory perception, compared to healthy controls. These neural network alterations were strongly linked to previously reported cognitive deficiency in FAP patients (Cruz-Correa et al., 2020). Observed differences in neural network cluster activation between FAP cases and healthy controls were correlated to cognition. Although this study cannot determine causality, this result indicates a strong association between cognition and brain-wide network activation. In combination with these cognitive deficits, the present data suggest that dysfunction in the APC protein affects functional neural connectivity in a broad range of cognitive and, perhaps, sensory brain networks and regions. Potential implications of the network specific findings and how they relate to previous literature in APC relevant topics, including depression, schizophrenia, autism, sensory perception and general cognition, are pertinent here.

The LFPN has been implicated as a network potentially involved in aphasias and in certain stroke patient’s decrease in language function (Zhu et al., 2014). The LFPN component identified in our analysis was primarily comprised of activation in the superior temporal gyrus, middle temporal gyrus, and precentral gyrus. Cognitive processing speed has been specifically linked to lateralization in FP networks (Chechlacz, Gillebert, Vangkilde, Petersen, & Humphreys, 2015). From an evolutionary and developmental perspective, specific and minor neuroanatomical changes in frontoparietal networks have substantial effects on cognition (Vendetti & Bunge, 2014), and developmental alterations of frontoparietal connectivity predict symptom load in high functional autism patients (Lin et al., 2019). It will be valuable to follow cognitive aging in FAP patients, as cognitive processing networks, including frontoparietal, are particularly susceptible to age related decline (Nashiro, Sakaki, Braskie, & Mather, 2017).

The Medial Visual Network (MVN) is essential for performing both simple and higher order visual tasks (Heine et al., 2012). This network presents as activity localized in a bi-temporal manner in the most anterior portion of the brain, centralized in the striatum (Castellazzi et al., 2014; Coppen, Grond, Hafkemeijer, Barkey Wolf, & Roos, 2018). Decreased connectivity in the MVN in schizophrenia and bipolar patients are associated with cognitive deficits (Jimenez, Riedel, Lee, Reavis, & Green, 2019). Furthermore, congenital hypertrophy of the retinal pigment epithelium, which could affect connectivity patterns in visual networks, has been observed in FAP (Laghmari & Lezrek, 2014; Traboulsi, 2005), although this was not examined in the present study. Decreased connectivity with the MVN was reported in temporal lobe epilepsy patients with cognitive deficits (H. Yang et al., 2018). It is postulated that connectivity alterations in the MVN adversely affect the processing of visual stimuli in FAP patients, resulting in decreases in processing speed and cognitive efficiency.

The Default Mode Network (DMN), used as a standard network in most neuroimaging studies, with its activity observed across a wide variety of tasks and pathologies (Haatveit et al., 2016), was also identified in our ICA analysis of FAP patients. This network is associated with autobiographical thought, including moments of self-reflection, and often deactivates when subjects are actively completing functional paradigms (Smith et al., 2009). Gray matter volume in regions typically showing activation resembling the DMN is predictive of the progression of mild cognitive impairment (MCI) to Alzheimer’s disease (AD)(Eyler et al., 2019; Petrella, Sheldon, Prince, Calhoun, & Doraiswamy, 2011; Wang et al., 2013). Late life depression is also associated with alterations in DMN connectivity (Alexopoulos et al., 2012), and the beneficial effects of mindfulness based cognitive therapy on depression symptoms (included cognitive deficits) are correlated with changes in DMN connectivity (Cernasov et al., 2019). In schizophrenia, deficits in typical activation of the prefrontal cortex (PFC) is related to specific components of the DMN (Hu et al., 2017), suggesting that future internetwork and whole brain connectivity analyses in FAP would be valuable (Pan et al., 2018; Varangis, Razlighi, Habeck, Fisher, & Stern, 2019).

The Task Positive Network (TPN) is generally comprised of two bilateral crescent-like clusters spanning the dorsolateral prefrontal cortex and sensorimotor areas around the temporoparietal junction (Corbetta, Kincade, & Shulman, 2002; Fox et al., 2005; Grady et al., 2010). This network is most active during attention demanding tasks and exhibits correlated fluctuations in activity with task accuracy. The TPN is commonly paired with the DMN as it is generally anticorrelated with the DMN activation, however, the DMN has similarly been shown to be active in specific tasks as well (Haatveit et al, 2016). Changes in TPN interaction with the DMN have been implicated in adverse cognitive patterns in depression, such as rumination (Hamilton et al., 2011). In a study of the performance of young and old participants in visual cognitive tasks, changes linked to decreased neurocognitive efficiency were observed in both the DMN and TPN (Grady et al., 2010). The TPN is also altered in patients with mild cognitive impairments (MCI) supporting the hypothesis that increased TPN activity may be a compensatory response to neurodegeneration (Melrose et al., 2018). Changes in the TPN and/or how it interacts with the DMN may mediate attention related aspects of cognitive deficits in FAP patients.

The major regions of the salience network (SN) include the amygdala, substantia nigra, thalamus, and hypothalamus (Seeley et al., 2007). This network plays a role in processing of rewards, cognitive control, and behavior (Ham, Leff, de Boissezon, Joffe, & Sharp, 2013). The SN is activated in the presence of particularly salient stimuli, and this network may be correlated with feelings of anxiety when assessed pre-MRI scan (Seeley et al., 2007). Typical internetwork connectivity patterns with the SN are disrupted in MCI, and the degree of disruption is associated with the degree of overall cognitive deficit (Chand, Wu, Hajjar, & Qiu, 2017). Similar to studies of the TPN, this role of SN in cognition was not limited to MCI, and the SN may be another region with a key role in age related cognitive changes (La Corte et al., 2016). Hyperconnectivity within the SN may be a defining feature of autism spectrum disorders (Uddin et al., 2013), and atypical connectivity between the DMN and SN suggest similar neural mechanisms in ASD and schizophrenia (Chen et al., 2017). While additional longitudinal data are needed, FAP mediated changes in the SN may be the result of abnormal neurodevelopmental and/or premature age related alterations.

The lateral visual network (LVN) has been attributed to helping to facilitate sensorimotor processes (Castellazzi et al., 2014). It commonly presents as activation in the occipito-temporal and parietal regions and is associated with the observation of highly appealing visual stimuli (Belfi et al., 2019). A study of late life depression, which often involves cognitive deficits, reported significant alterations in visual networks (Eyre et al., 2016). In addition, both major depression and schizophrenia patients exhibit differences in visual network connectivity compared to healthy controls (Wu et al., 2017). Hence, FAP related changes in both the lateral and medial visual networks may adversely affect the processing and response to visual stimuli.

The Executive Control Network (ECN) is comprised of fronto-localized activation, with regions including the anterior cingulate and paracingulate (Zhao, Swati, Metmer, Sang, & Lu, 2019). This network is important in facilitating executive function. One study examining cirrhotic patients (liver disease) showed that patients performed significantly worse than controls on a Stroop Task as well as the performance showing a significant correlation with disrupted ECN connectivity (Yang, Chen, Chen, & Lin, 2018). Similarly, this network has recently been implicated in studies of major depressive disorder, with increased activation to specific network-related areas (Zhao et al., 2019). A lack of segregation between ECN and DMN activation was correlated with decreased processing speed in a longitudinal study of healthy older adults (Ng, Lo, Lim, Chee, & Zhou, 2016). As individuals age, the functional specialization of these two networks is attenuated. Decreased network segregation is implicated in major depression, where connectivity with the ECN and between the ECN and DMN is disrupted compared to non-depressed controls, potentially representing compensatory cognitive mechanisms (Albert, Potter, Boyd, Kang, & Taylor, 2019). Future studies of the mechanisms mediating cognitive deficits in FAP patients should focus on the role of the ECN, its functional connections with the DMN, and potential moderating factors, such as depression.

The auditory network presents as activation in the insula, auditory cortices, cingulate, and occipital cortices (Maudoux et al., 2012), and is primarily responsible for processing and parsing auditory stimuli. This network has been investigated for its potential involvement in processing fearful stimuli, with emotional “hubs” being associated with its activity (Koelsch, Skouras, & Lohmann, 2018). Auditory deficits have been reported in FAP patients in the absence of significant differences in IQ (O’Malley et al., 2010), although another investigation did not observe significant impairments (Jones et al., 2010). The APC protein is essential for cellular processes related to cochlear sensitivity, and since these are common cellular processes, it is suggested that APC mediates synaptic maturation in wide variety of tissues (Hickman, Liberman, & Jacob, 2015). It is possible that auditory deficits in FAP patients could be mechanistically mediated by effects at the ear and/or auditory processing nuclei.

### Limitations

Main limitations of this study include the use of an aggregate cognitive score due to small sample size, the presence of only one imaging time point, and the lack of structural data. While the use of an aggregate cognitive score precludes associations between specific types of cognition and network connectivity changes, this was justified based on the consistency across cognitive scores, the small sample size, and the focus on general conclusions strongly supported by the imaging data. Future studies would benefit from additional data points and the inclusion of structural MRI data to investigate the etiology of these cognitive deficits in FAP patients in early through late life. Although structural changes are likely given the role of APC in neurodevelopment and structural-functional relationships in cognition, the current manuscript can only speculate on these changes due to the focus on functional data.

### Conclusion

Resting state functional connectivity in FAP patients is substantially disrupted in a broad range of networks critical to cognition and visual and auditory stimuli processing. These alterations in connectivity are strongly associated with overall cognitive deficits in these individuals. Taken together, the APC gene appears to play a critical role in neurocognitive function and sensory processing, and could negatively affect development through changes in neural connectivity. Future studies should characterize changes in neurocognition and auditory and visual sensation and processing throughout the lifespan to characterize the progression of related deficits in FAP patients. The role of the APC protein in other neurodegenerative disorders, such as autism and schizophrenia, is warranted.

## Data Availability

Data will be made available via the Open Science Framework.

## Financial Disclosure

The authors have no financial conflicts of interest to report.

## Funding

Supported by Award Number 2U54MD007587 from the National Institute on Minority Health and Health Disparities and National Institute of Allergy and Infectious Diseases and the Investigational Fund of the University of Puerto Rico Vice Presidency for Research.

## Notes

### Competing Interest Statement

The authors have declared no competing interest.

